# The Future is Green. Integrating Green and Blue Space Data from European Urban Atlas into UK Biobank

**DOI:** 10.1101/2022.05.09.22274764

**Authors:** Mariya Geneshka, Colin J McClean, Simon Gilbody, Joana Cruz, Peter Coventry

**Affiliations:** Department of Health Sciences, University of York, York YO10 4DD, UK; Department of Environment and Geography, University of York, York, YO10 5NG, UK; Hull York Medical School, University of York, York, YO10 5DD, UK; Population, Policy and Practice Department, UCL Great Ormond St. Institute of Child Health, London WC1N 1EH, UK; York Environmental Sustainability Institute, University of York, York YO10 4DD, UK

**Author notes:** Corresponding author(s). Department of Health Sciences, University of York, York YO10 4DD, UK.

**Keywords:** environmental epidemiology, methods, data linkage, data manipulation, green space, blue space, large cohort, health, chronic health, evidence-based research

## Abstract

**Background:** Green and blue spaces can promote good physical and mental health and prevent the development of long-term conditions. Evidence suggests that not all green spaces affect health equally, and that certain types and properties of green spaces are stronger predictors of health than others. However, research into the causal mechanisms is limited in large cohorts due to lack of objective and comparable data on green space type, accessibility, and usage.

**Methods:** We used data from Urban Atlas to compute measures of urban park accessibility, street trees availability, and total green and blue space availability for 300,000 UK Biobank participants. Exposure metrics were computed using circular buffers with radii of 100 m to 3000 m. Pearson correlation coefficients and other descriptive statistical parameters were used to test agreement between variables and explore the utility of indictors in capturing different types of green spaces.

**Results:** Strong positive correlations were observed between variables of the same indicator with different buffer sizes. The presence of park and proportion of street tree canopy variables were negatively correlated with amount of total green space variables. This signifies distinct differences in type of green spaces captured by these variables.

**Conclusions:** Overall, five distinct indicators of park accessibility, street trees availability, and total green and blue space availability have been integrated into a large sample of the UK Biobank. Our method is replicable to settings across Europe and facilitates evidence-based research on the roles of different green and blue spaces in health promotion and ill-health prevention.

**Key Messages:**

- Different types of green spaces and their position in the neighbourhood can promote and protect health by mitigating pollution and increasing physical activity and socialisation.
- We present the methods of constructing and linking data on urban green spaces, street trees and natural vegetation into a large health cohort, the UK Biobank.
- The ability to distinguish between types of green spaces and their intended use can help inform public health interventions, influence urban policy, and aid urban planning in building sustainable and healthy cities.
- Our methods are transferable and will allow others to explore the links between environment and health in UK Biobank and other health cohorts.

## Introduction

Green and blue spaces are umbrella terms used to describe the presence of vegetation (green) and water (blue) bodies in the surrounding environment.[1] Green spaces can affect health through several bio-physiological pathways, including promoting health-related behaviours like physical activity; increasing socialisation and connectedness with people and nature; and reducing the presence of air pollutants and noise.[2, 3] Exposure to green space can potentially reduce the risk of several non-communicable diseases (NCDs). In a longitudinal study, for example, the risk of developing cardio-vascular disease (CVD), myocardial infraction, and stroke was respectively 15%, 23% and 13% lower in those who had more green spaces in their neighbourhood compared to those who had fewer green spaces.[4] Moreover, meta-analyses found 28% reduction in the risk of type II diabetes and 23% reduction in the risk of stroke mortality in those with greater amount of green space in their neighbourhood.[5, 6]

Green spaces also impact mental health and wellbeing. Observational, epidemiological studies found higher amount of green space was associated with lower odds of stress, higher self-perceived general health, and higher quality of life.[7-9] Furthermore, longitudinal research showed that those living in areas with lowest amount of residential greenery had 24% to 52% higher risk of developing schizophrenia compared with those living in areas with most greenery.[10, 11] The relationship between green space and depression is less well understood, but cross-sectional studies showed a moderate decrease in the odds of depression with greater amount of green space and higher accessibility to parks of large areas.[12, 13] Growing evidence suggests that, in addition to amount of green space, different types of green and blue space environments affect health differently.[14] Green areas, such as grasslands, serene environments, higher number of forests and higher number of urban green spaces reduced the risk of poor mental health.[15, 16] However, no protective relationships with mental health were observed for exposures to saltwater bodies, wetlands, rangeland, and agricultural land.[15, 16] In urban settings, higher availability of street trees, but not higher availability of grass or total green space, was associated with lower odds of diabetes and CVD events.[17, 18] Furthermore, a comparison between higher availability of public parks and total green space area showed that public parks reduced blood pressure while larger areas of green space in the neighbourhood showed no significant relationship with blood pressure change.[19]

Type, position, and duration of exposure to green/ blue space affect health at different rates and potentially through different causal pathways. In systematic reviews, higher amount of street trees, good accessibility to parks and some types of land use classes showed stronger relationships with health than higher availability of grass or total green space.[20-22] However, the mechanisms and direction of causation behind these relationships are still not fully understood. This is partly due to lack of high quality comparable observational research. Despite the World Health Organisation’s (WHO) [23] call to include more objective and comparable measures of green space accessibility and usage in evidence-based research, cohort studies still mainly use single measures of greenery, such as the Normalized Difference Vegetation Index (NDVI), or proportion of green space.[13, 14, 24] Reasons for this include lack of availability of objective environmental data in health cohorts, and limited characterisation of different green spaces present in the surrounding environment.[24]

The UK Biobank is a comprehensive resource for studying the complex interactions between the environment and health. However, studies on the relationship of exposure to green and blue spaces and health have mainly used NDVI and data from older land use datasets to capture proportion of greenery on limited spatial scales.[25-28] We describe here the methods to link high resolution data on street trees, parks, and amount of green and blue space from 2006 and 2012 Urban Atlas dataset into the UK Biobank. Based on prior research, we hypothesise that it is certain types of green spaces, such as parks and street trees, and their properties (e.g., type, position, size, or intended use) that influence health through different bio-physiological pathways (fig. 1).[21, 22] Therefore, the aim of this study is to construct multiple distinct indicators of green and blue space availability and accessibility that can be used in evidence-based health research. To achieve this, we first describe the process and methods of computing and integrating variables of urban park accessibility, street trees availability, and total green and blue space availability into UK Biobank participants using geographical information systems programming. Second, we calculate the statistical parameters of these variables to descriptively characterise each exposure metric and its spatial scale. Third, we use correlation coefficients to test the agreement between variables and explore the ability of indicators in capturing different types of green spaces in the surrounding neighbourhood. This linked dataset will be used to model the cross-sectional associations between green space availability and accessibility and the prevalence of multimorbidity disease clusters. It will also become available for anyone seeking to conduct research using UK Biobank data.

**Figure 1:**
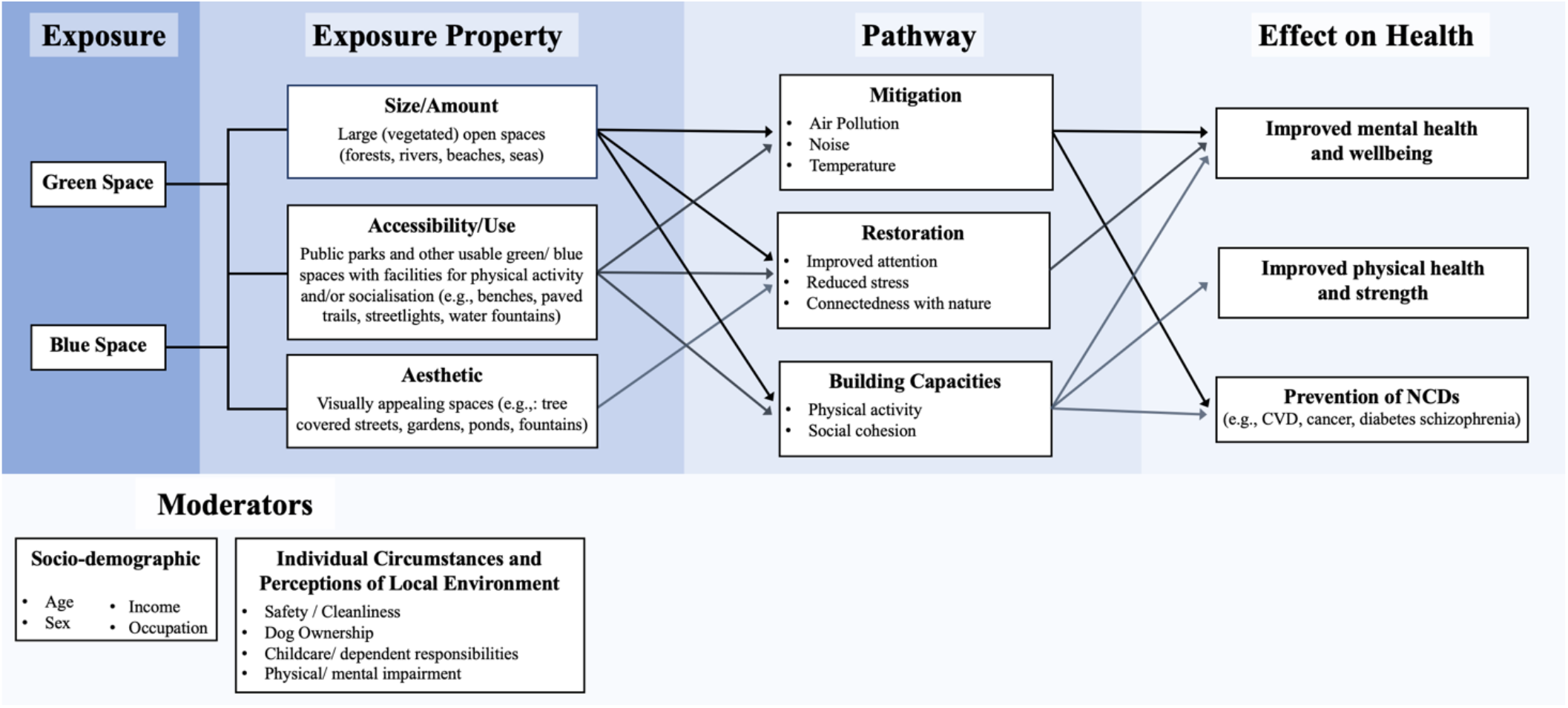
Conceptual framework for the relationship between green and blue space properties and health

## Methods

### Description of Data Sources

#### UK Biobank

The UK Biobank is a large, population-based prospective cohort of 502,650 men and women aged 40-69 years.[29] Recruitment for voluntary participation started in 2006 and finished in 2010. Participants were selected if they resided within 25 miles of one of the 22 UK Biobank assessment centres around the UK and were registered with a National Health Service (NHS) General Practitioner (GP). Data about participants’ socio-demographic characteristics, living arrangements, lifestyle factors, built-environment exposures, and medical history were collected through touchscreen questionnaires and nurse-led interviews. Follow-up of the cohort is ongoing and disease status and mortality are tracked through electronic health records and cancer registries.[29]

#### Urban Atlas

The European Urban Atlas (UA) is a land use dataset covering over 200 European Functional Urban Areas (FUA) with a population of at least 50,000 people.[30, 31] FUAs are defined by the European audit as the area of a metropolitan city and its surrounding commuting zone. The commuting zone is the area around the city where at least 15% of the employed residents are commuters into the city.[32] The UA dataset contains nomenclature of 20 land use classes, 17 of which are built-environment classes with a minimum mapping unit (MMU) of 0.25 ha and 3 are natural classes with a MMU of 1 ha. The overall minimum accuracy for the data is 80% and the minimum mapping width is 10 m. The UA datasets are collated from topographic maps, 2.5 m Earth Observation spatial resolution multispectral data, SPOT 5 satellite imagery and other Very High Resolution (VHR) imagery for the years 2006, 2012 and 2018.[31] The data is open access and available from the European Environment Agency and the Copernicus Land Monitoring website (https://land.copernicus.eu).

The validity of UA in capturing amount of green space has been previously tested against the CORINE, UK Land Cover Map and NDVI datasets. It was established that UA produces comparable results to all three of these indicators.[33, 34] The *Urban green areas* layer from the UA has also been endorsed by WHO [23] as a suitable indicator of urban green space accessibility and is a preferred indicator for capturing usable green spaces in urban areas due to its high resolution and ability to measure green space change over time.[35]

### Data Selection and Processing

UK Biobank participants’ residential address location coordinates (rounded to 100 m accuracy) at baseline were used as proxies for permanent residence. UK Biobank participants predominantly live in urban areas and UA data is available for about 300,000 UK Biobank participants who reside within the boundaries of the following FUAs: London, Bristol, Cardiff, Stoke-on-Trent, Nottingham, Sheffield, Leeds, Manchester, Liverpool, Newcastle upon Tyne, Edinburgh, and Glasgow.

### Exposure Metrics

Several natural and artificial land use classes were used to compute exposure metrics of accessibility to park; availability of street tree canopy; availability of total green space; availability of total blue space; and availability of total green and blue space at different spatial scales (see table 1 for further information).[36]

**Table 1:** Descriptions of Computed Exposure Metrics

| Exposure Indicator | Exposure Metric | UA Nomenclature used (year of data collection) [36] | UA Nomenclature Description [40] | Buffer Sizes |
| --- | --- | --- | --- | --- |
| Accessibility to Usable Urban Park | Distance to Park | 14100 Green urban areas (2006) | Green areas accessible to the public for predominantly recreational use, such as public parks, gardens, and zoos. | N/A |
|  | Presence of Park |  |  | 300m, 1500m |
| Availability of Street Trees | Amount of Street Tree Canopy | Street Tree Layer (2012) | Contiguous patches of trees ($\geq 5$ m height) covering an area of at least 500m<sup>2</sup> over Level 1 *Artificial surfaces* nomenclature layers | 300m, 1500m |
| Availability of Total Green Space | Amount of Total Green Space | 14100 Green urban areas (2006)<br>20000 Agricultural + seminatural + wetland areas (2006)<br>31000 Forests (2006) | *Agricultural + seminatural + wetland areas* layer: range of natural open spaces including arable land, pastures, grasslands, arable crops, moors, beaches, bare rocks, snow and ice and wetlands.<br>*Forests* layer: areas with tree canopy coverage of over 30% and tree height of over 5 m. | 100m, 300m, 1500m, 3000m |
| **Availability of Blue Space** | Amount of Total Blue Space | 5000 Water Bodies | Visible water surface area of rivers, lakes, ponds and canals | 100m, 300m, 1500m, 3000m |
| **Availability of Total Green and Blue Space** | Amount of Total Green & Blue Space | 14100 Green urban areas (2006)<br>20000 Agricultural + seminatural + wetland areas (2006)<br>31000 Forests (2006)<br>5000 Water Bodies | See above | 100m, 300m, 1500m, 3000m |

Accessibility to urban park was measured as the straight-line (Euclidean) distance from a participant’s residential address to their nearest public park using data from the UA 2006 nomenclature layer: *Green urban areas*.

Availability of street trees in the neighbourhood was measured as amount (proportion) of street tree canopy in straight-line (Euclidean) radial distance buffers around the residential address. Availability of total green space, blue space and total green and blue space was measured in amount (proportion) within a straight-line (Euclidean) radial distance buffers around the residential address. The UA data layers used to construct each indicator are described in Table 1.

### Data Processing and Analysis

All data were processed in ArcGIS Pro, a 64-bit Desktop application facilitated by Esri ArcGIS Platform. The 2006 UA vector datasets were rasterized to 50 m resolution and the 2012 *Street Tree Layer* dataset was rasterised to 10 m resolution. The six-digit residential location coordinates of UK Biobank participants were overlayed with the rasterized UA data. Proportion green/blue space within distance buffers and the Euclidean distance to park were then computed for each UK Biobank participant (see table 1 for further details on buffer size). Participants whose residential address or buffer area fell on or outside the boundary of the UA data were excluded.

### Data Analysis

The derived environment data was transferred from ArcGIS Pro to RStudio for analysis. Descriptive statistics, such as central tendency (i.e., means, medians), frequencies and dispersion measures (i.e., standard deviation and inter-quartile ranges) were calculated. Bivariate data analysis, chi-squared tests, Pearson tetrachoric, point biserial and product moment correlations were also applied according to data type.

## Results

Table 2 describes the statistical parameters of the computed green and blue space variables. Just under two-thirds (62.94%) of UK Biobank participants had a park within 300 m of their residential address and almost all (98%) had a park within 1500 m of the residential address. The mean straight-line distance to a park from the residential address was 291.48 m (table 2). The Chi-squared test suggests there’s strong correlation between presence of park in 300 m and presence of park in 1500 m variables.

**Table 2:** Statistical parameters of computed green and blue space variables

| **Metrics** | **Var. Name** | **n**<br>**(observed**<br>**cases)** | **Mean**<br>**(%)** | **Standard**<br>**Deviation** | **Median**<br>**(%)** | **Min**<br>**(%)** | **Max**<br>**(%)** |
| --- | --- | --- | --- | --- | --- | --- | --- |
| Total green space in 100m | total_100_green | 311,451 | 10.26 | 20.32 | 0.00 | 0.00 | 100.00 |
| Total green space in 300m | total_300_green | 308,979 | 15.40 | 18.32 | 8.84 | 0.00 | 99.03 |
| Total green space in 1500m | total_1500_green | 239,747 | 27.78 | 20.57 | 21.19 | 0.04 | 99.74 |
| Total green space in 3000m | total_3000_green | 192,094 | 32.31 | 21.10 | 25.94 | 1.47 | 98.85 |
| Total blue space in 100m | blue_100 | 311,451 | 0.24 | 2.68 | 0.00 | 0.00 | 95.49 |
| Total blue space in 300m | blue_300 | 308,979 | 0.46 | 2.66 | 0.00 | 0.00 | 87.54 |
| Total blue space in 1500m | blue_1500 | 271,118 | 1.12 | 2.55 | 0.21 | 0.00 | 50.51 |
| Total blue space in 3000m | blue_3000 | 219,462 | 1.20 | 1.70 | 0.56 | 0.00 | 47.92 |
| Tree canopy in 300m | tree_300 | 138,831 | 20.03 | 17.13 | 15.30 | 0.04 | 99.60 |
| Tree canopy in 1500m | tree_1500 | 171,734 | 19.14 | 13.47 | 16.70 | 0.00 | 85.06 |
| Total green and blue space in 100m | green_blue_100 | 311,451 | 10.48 | 20.51 | 0.00 | 0.00 | 100.00 |
| Total green and blue space in 300m | green_blue_300 | 308,979 | 15.86 | 18.52 | 8.84 | 0.00 | 100.00 |
| Total green and blue space in 1500m | green_blue_1500 | 239,719 | 28.93 | 20.50 | 22.85 | 0.29 | 99.74 |
| Total green and blue space in 3000m | green_blue_3000 | 180,572 | 34.04 | 21.19 | 27.70 | 2.10 | 99.02 |
| Distance to park | distance_park | 312,284 | 291.48* | 378.90* | 201.15* | 0.00* | 15843.71* |
|  | Var. Name | Yes (%) |  |  | No (%) |  |  |
| Presence of park in 300m | park_300 | 194,481 (62.94%) |  |  | 114,498 (37.06%) |  |  |
| Presence of park in 1500m | park_1500 | 273,172 (98.14%) |  |  | 5,190 (1.86%) |  |  |
| X-squared = 9902.8, df = 1, p-value < 2.2e-16 |  |  |  |  |  |  |  |

\* Unit of measurement is meters (not percentage)

Median and interquartile range values for all other environment variables are shown in box and whisker plots in figure 2. Overall, the data are skewed towards the null. The median amount of green/blue space increases with buffer size. The median amount of total green space in a 100 m buffer is around 0%. This increases to 26% in a 3000 m buffer. Median amount of blue space follows a similar pattern, but values tend to stay around 0% for all buffer sizes. Median amount of street tree canopy is 15% in a 300 m buffer and 17% in a 1500 m buffer.

**Figure 2:**
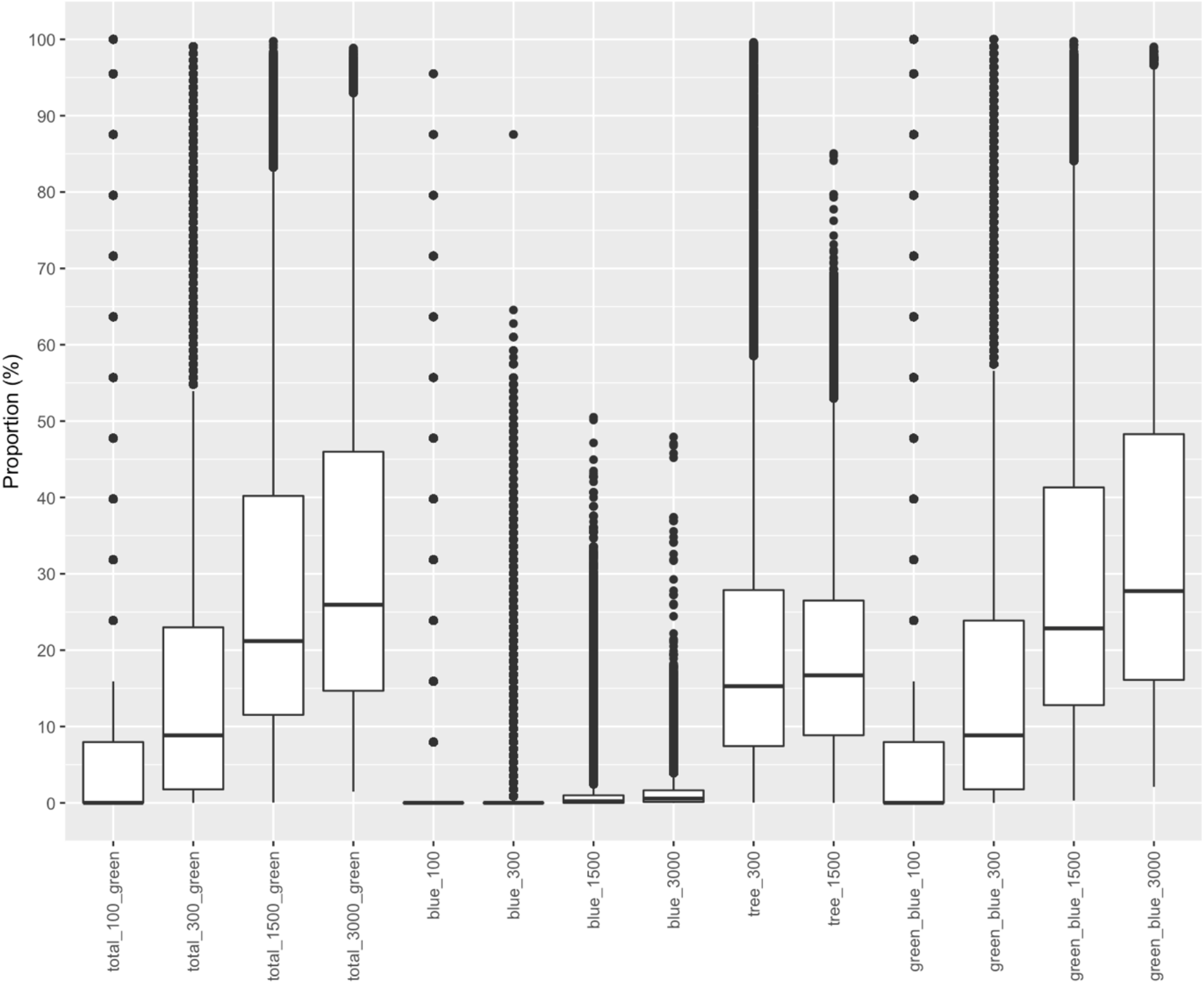
Box and whisker plots of variables

Figure 3 shows a Pearson correlation matrix of the computed variables. Overall, variables of the same exposure indicator with different buffer sizes tend to have strong positive correlations with each other. The correlation coefficients are weaker between variables that have larger buffer size differences. There’s strong positive correlation between street tree canopy in 300 m buffer and street tree canopy in 1500 m buffer (r = 0.78).

**Figure 3:**
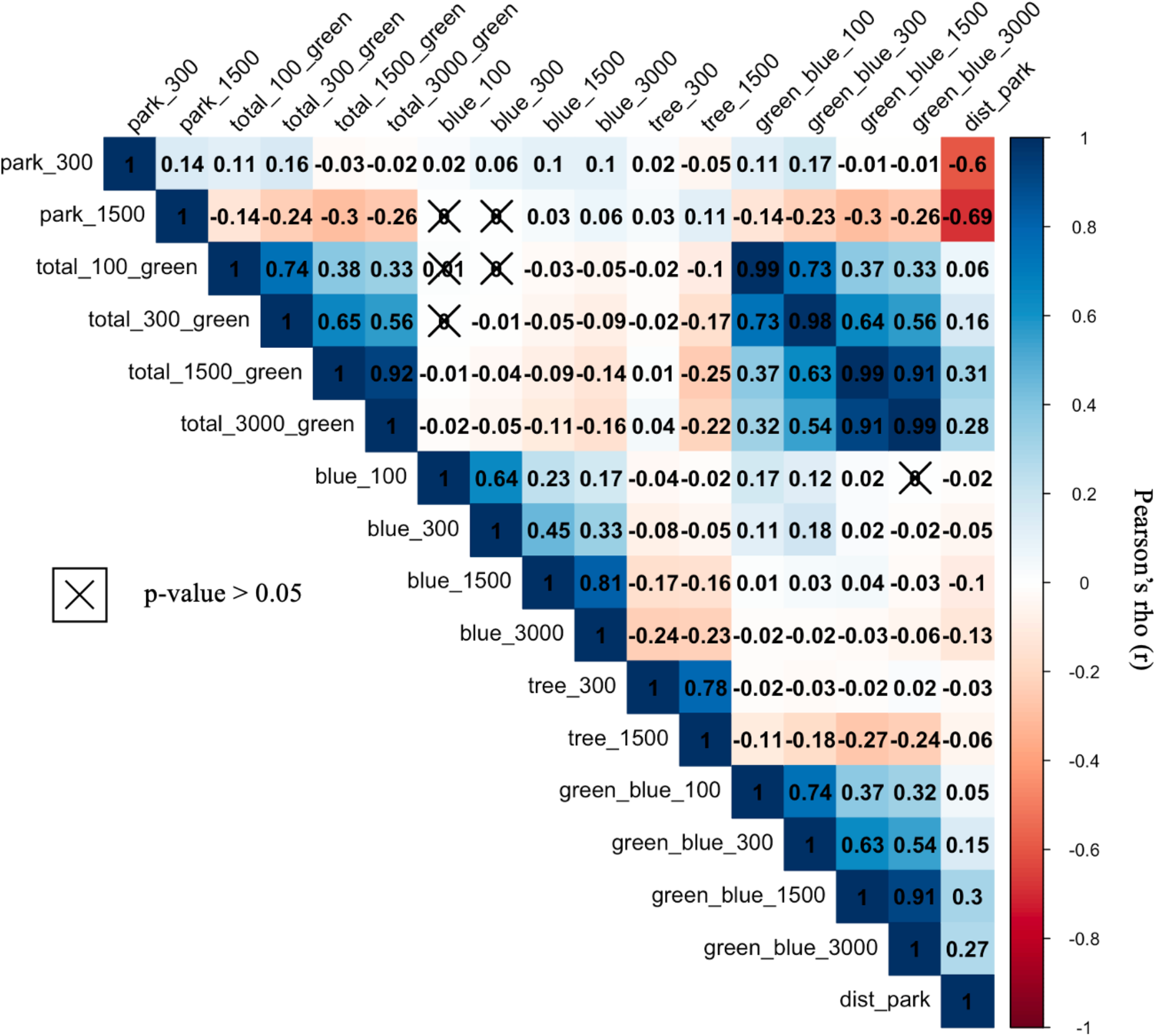
Pearson correlation matrix of variables

The correlation matrix also shows negative correlations between the presence of park variables, street tree canopy variables and amount of total green space variables. Weak positive correlations were observed for presence of park in 300 m buffer variable (park_300) with amount of total green space and amount of total green and blue space variables. However, weak to moderate negative correlations were observed for presence of park in a 1500 m buffer variable (park_1500) with amount of total green space and amount of total green and blue space variables (fig. 2). There are also weak negative correlations between the amount of street tree canopy in 1500 m buffer variable (tree_1500) and the amount of total green and blue space variables.

## Discussion

### Interpretation of Computed Green Space Indicators

In this data linkage study, we present the process and results of linking green and blue space exposure data from Urban Atlas with the UK Biobank. Five distinct indicators were constructed: accessibility to park; availability of street trees; availability of total amount of green space; availability of total amount of blue space; and availability of total amount of green and blue space. Accessibility to park is measured by the presence of a park in 300 m buffer, presence of park in 1500 m buffer, and the Euclidean distance to nearest park. Having a park within 1500 m of the residential address was negatively correlated with having higher amount of total green space. The amount of total green space variables were computed using two natural land use layers: *Forests* and *Agricultural + seminatural + wetland areas*, and one artificial surfaces layer: *Green urban areas*. The *Green urban areas* layer was used to compute the presence of park variables and while there’s overlap in data, the negative correlations between the two indicators suggest that participants who have high proportion of total green space in 1500 m buffer around their residential address likely live on the rural-urban fringe and have higher availability of forests and agricultural vegetation rather than urban green areas. The amount of total green space variables, therefore, are indicators of availability of naturally growing outdoor vegetation, while the presence of park variables are indicators of accessibility to usable urban green spaces.

With the emerging evidence on the health-promoting role of street trees we also aimed to compute an indicator of street trees availability.[20] This is captured by the amount of street tree canopy variables. Higher proportion of street trees showed to be negatively correlated with both higher proportion of total green space and presence of parks. This is expected because UA nomenclature, *Street Tree Layer*, is a non-overlapping dataset of patches of street tree canopy over artificial surfaces only.[36] These results show that, together, the five indicators are distinct but complementary to each other. They each capture different types of green spaces in the surrounding neighbourhood. The street trees and distance to park variables are indicators of two types of accessible green spaces: public parks and trees lining roads and paths. The amount of total green space variables, on the other hand, are measures of natural vegetation mainly found on the fringe of rural and urban areas.

### Implications for Health Research and Policy

The integration of multiple, distinct green space indicators into UK Biobank addresses an important data gap in health research. Lack of high-quality, comparable data on green space accessibility, availability and use in epidemiological studies and in the UK Biobank has resulted in little research into the causal mechanisms behind the relationship of exposure to green and blue space and health.[24, 37, 38] The ability to distinguish between types of green spaces, their position in the neighbourhood and their intended use can strengthen evidence-based research and inform public health practice. Outdoor green and blue spaces can be utilised to deliver targeted interventions to promote healthy lifestyles and prevent ill-health. This research can also help wider urban planning and policy in creating heathier urban environments where green and blue spaces are integral parts of healthy daily living.[23]

### Strengths and Limitations

This data linkage study has several strengths. To the best of our knowledge, it is the first study to integrate multiple comparable indicators of urban park accessibility and street trees availability into a large, population-based UK cohort. Previously, data linkages have been limited to single area case studies or focused on built-environment indicators.[24] The large sample size of the UK Biobank improves precision and power of future research, avoids bias, and opens opportunities to explore the still limited but strong evidence for the health-promoting benefits of street trees, parks and natural vegetation.[39-41] Furthermore, our method and exposure metrics are replicable to settings across Europe. The UA dataset covers urban areas across Eastern and Western Europe, allowing objective exposure assessment for different populations and settings.[36]

We also computed exposures at an individual level, which means every participant has a unique measure of greenery in their neighbourhood. This improves accuracy in exposure measurement and prevents ecological bias (and potential fallacy) that commonly occurs when aggregate measures are used in individual analyses.[42] The computation of buffer sizes ranging from 100 m to 3000 m, on the other hand, allows for comparative research at different spatial scales. Three-hundred meters is recommended as the maximum distance anyone should live from an accessible green space; however, this figure is arbitrary, and research has shown that both small (500 m) and large (up to 2000 m) buffer sizes can be strong predictors of health. [23, 43, 44]

Our data linkage study has multiple strengths, but it is not without limitations. Our analyses are primarily based on measuring proportion of green and blue space within straight-line distance buffers. These buffers capture any greenery that falls within the buffer’s area, irrespective of the green space’s accessibility or ownership. One advance in geo-computation has been the ability to capture greenery only along publicly accessible roads and paths.[45] This approach might be useful for some exposure metrics, such as street trees, but it’s also computationally intensive and doesn’t necessarily improve accuracy in exposure measurement because circular and road-network buffers tend to capture similar amount of greenery.[46]

Other limitations include the geo-processing resolution of the data. The 2006 UA layers were converted to raster datasets with a resolution of 50 meters. Rasterising a vector feature may cause potential data loss of areas smaller than the size of the raster grid cell. We chose 50 meters as a grid cell size because it facilitated geoprocessing for large sample sizes. Smaller grid cell sizes, while more accurate, could not allow large scale data analysis due to large memory storage.

## Conclusions

In conclusion, we have described a unique set of methods and results about how to link high quality, open-access land use data from Urban Atlas with the UK Biobank health cohort. We produced five novel indicators of urban park accessibility; street tree availability; and total green/ blue space availability. This broadens opportunities for comparable epidemiological research in a large health cohort. Given the availability of UA data for UK and Europe, we believe our methods are also replicable to settings, populations and curated cohorts outside of the UK Biobank.

## Data Availability

Data fields derived from this study and all other data underlying this article is owned by UK Biobank and can be accessed with an approved project application from the UK Biobank's Access Management System (AMS). Raw environment data from Urban Atlas is open access and available from European Environment Agency.

## Ethics approval

Researchers do not require a separate ethics approval to use UK Biobank data and are covered under the UK Biobank ethics approval by the North West Multi-centre Research Ethics Committee (MREC) as a Research Tissue Bank. This project was approved by the UK Biobank’s Access Management System (AMS), application no. 73700, and grants access to restricted UK Biobank Fields 22701 and 22703 (*home location – east coordinate* and *home location – north coordinate*).

## Author contributions

MG designed and led the development of the study. MG produced the data fields, conducted the data linkage and drafted the paper. PC, CM, JC and SG made critical intellectual contributions to the paper. CM advised and assisted on the geoprocessing of the data. JC sourced the UA data, formulated the theoretical background underpinning the green and blue space indicators construction and the metric spatial scales. PC formulated the theoretical background underpinning the study’s aims and objectives, and aided the interpretation of results in the discussion section. SG advanced the development of the introduction and discussion sections and helped obtain UK Biobank data. PC and SG obtained funding for the UK Biobank data. PC, CM and SG reviewed and edited the paper.

## Data availability

Data fields derived from this study and all other data underlying this article are owned by UK Biobank and can be accessed with an approved project application from the UK Biobank’s Access Management System (AMS). Raw environment data from Urban Atlas is open access and available from European Environment Agency.

## Supplementary data

None

## Acknowledgements

We’d like to thank Prof. Piran White from Department of Environment and Geography at University of York, Dr. Rachel Pateman from Stockholm Environment Institute, and Dr. Andre Bedendo De Souza from Health Sciences Department at University of York for their help and intellectual contribution to this paper. Their expertise in environmental data analysis and statistics has helped shape this project.

We’d also like to acknowledge the UK Biobank for providing the participant location coordinate data and storing and maintaining the derived data fields in this study. Finally, we’d also like to acknowledge the European Environment Agency for funding and facilitating the development of the Urban Atlas dataset. The Urban Atlas is an open access resource available from Copernicus Land Monitoring (**https://land.copernicus.eu)**.

